# Exploring pathways to compulsory detention in psychiatric hospital and ways to prevent repeat detentions; Service user perspectives

**DOI:** 10.1101/2024.06.04.24308425

**Authors:** Mary Birken, Ariana Kular, Patrick Nyikavaranda, Jordan Parkinson, Lizzie Mitchell, Kathleen Lindsay Fraser, Valerie Christina White, Janet Seale, Jackie Hardy, Cady Stone, Mark Keith Holden, Tyler Elliott, Zishi Li, Henrietta Mbeah-Bankas, Lisa Wood, Fiona Lobban, Brynmor Lloyd-Evans, Sonia Johnson

## Abstract

**Purpose:** This study, co-produced by a team of academics, lived experience researchers and clinicians, explores the views and experiences of people who have been compulsorily detained in hospital under the Mental Health Act (1983) (MHA) in England, to understand how and why, from their perspective, compulsory detentions occur, and what might help prevent them.

**Methods:** Semi-structured qualitative interviews were conducted with 20 people (55% male, 40% Black/Black British, 30% White British) who had been compulsory detained in hospital within the past 5 years. Lived experience researchers with relevant personal experience carried out interviews via telephone or videoconference and participated in analysis of data via a template approach.

**Results:** We derived three over-arching themes from interviews. The first theme “Individual factors increasing or reducing likelihood of being detained” encompassed factors related to people’s own lives and attitudes, including life stressors, not taking medication, the risk individuals may pose to themselves or others, and their attitude to and management of their mental health. The second theme “Family and Support Network” reflects how attitudes and support from family, friends and support network may contribute to compulsory detentions or support people to stay well. The third theme “Need for improvement in Service Responses” identified limitations of services that contribute to detention, including lack of collaborative care and choice, poor quality of professional support, and discriminatory attitudes from staff. Each theme also included potential approaches to addressing these limitations and reducing compulsory detentions.

**Conclusion:** Findings suggest multiple interacting factors may lead to people being detained in hospital under the MHA, and that improvements to services, such as increasing collaborative care and service user-led family involvement, could prevent further detentions.

## Introduction

Compulsory detention in mental health wards is increasing in some higher income countries, including England [1–3]. Reducing compulsory detentions in hospital is a national and international priority as it is often a distressing and traumatising experience for service users and their carers, and may lead to prolonged and disrupted recovery and poor therapeutic alliances [4–5]. Compulsory detentions are also expensive, recently estimated as costing £18,315, equating to approximately €20,100, or $23,000. per detention, diverting resources from longer term preventive and recovery-approaches that may have better-established positive impacts [6].

While service users report some positive outcomes from compulsory detentions [4], experiences appear predominantly negative [4, 7, 8]. Compulsory detention by its nature infringes principles of informed consent, collaboration and joint decision making otherwise deemed essential in mental health care [9]. Mitigating harms, optimising experiences and outcomes and preventing further detentions where possible is thus a high priority.

In the UK, Section 2 or 3 are the most frequently used provisions in the MHA (1983), allowing people to be detained in hospital for a specified period where it is deemed necessary for their own health or safety, or protection of others while the individual is assessed or when treatment cannot be given unless detained [10].

In England, there are large ethnic inequalities in risk of being detained under this legislation, with people from Black Caribbean and Black British backgrounds having an estimated four times higher risk than White British individuals [11–12]. Qualitative research has found that Black people who have been detained often feel that the decision to detain them and coercive or traumatic experiences in hospital are linked to racism [13]. Reducing risk of detention is thus especially important for these over-represented ethnic groups.

Currently, research evidence on how to prevent detentions and repeat detentions remains very restricted, with some evidence suggesting crisis planning-based interventions may reduce repeat detentions, but little other positive evidence to build on [14, 15]. An enhanced understanding of risk factors for, and pathways to, compulsory detention has potential to inform interventions to reduce detentions. Quantitative studies yield some evidence on what is associated with greater risk of compulsory detention. Systematic reviews of risk factors for detentions among adults have identified diagnosis of a psychotic illness, previous episodes of involuntary hospitalisation, belonging to an ethnic minority, especially Black British, Caribbean and African groups, male gender, unemployment, single marital status, not owning a home, and receiving state benefits as risk factors [11, 16]. A recent narrative review [17] focuses on quantitative evidence regarding contextual and societal factors that may be drivers of rates of compulsory admission, including limitations in services, such as lack of alternatives to admission, and societal factors such as austerity and high unemployment. Most of the literature included in reviews focuses on compulsory detention in general rather than repeat compulsory admission, but an investigation of repeat compulsory admission in the Netherlands found that previous history of mental health treatment or homelessness and poor self-care were among risk factors [18]. Awareness of risk factors is useful for development of preventive interventions in that groups at high risk of detention can be focused on, but quantitative evidence so far yields only limited explanations of mechanisms underpinning these risk factors and pathways to compulsory admission [19, 20].

Qualitative research has potential to contribute to understanding of risk factors for and pathways to compulsory detention from the point of view of those detained, as well as families and clinicians. However, qualitative investigations of experiences of compulsory detention have tended to focus on experiences of being a detained inpatient [4] or on decision making at the point of admission [21] rather than on perceived reasons for and pathways to detention. Participants interviewed in a Norwegian study using grounded theory suggested that medicalising mental illness rather than considering difficulties in the context of people’s lives contributed to involuntary detention, with a lack of focus on addressing the social difficulties in people’s lives seen as contributing to mental health problems and to the crisis resulting in detention [22]. The study also reported that doctors did not have enough time during consultations to properly explore the patient’s circumstances. Further qualitative research exploring perceived reasons for involuntary detention has potential to contribute to development of approaches to reduce future compulsory detentions.

The present study is part of the first phase of an NIHR (National Institute of Health and Social Care research) funded study, the FINCH study (NIHR201739). The aim of the FINCH study is to adapt an existing intervention based on crisis planning that showed promise in reducing compulsory detentions in a study in Switzerland [23], for use in the UK, and to test it in a feasibility trial. Our findings from the current study informed the adaptation of the intervention by a co-production group including service users, clinicians and researchers. We describe the intervention development process and the trial protocol in a separate paper [24]. By interviewing people who had been compulsorily detained in England in the past 5 years, we aimed to understand how and why, from their perspective, compulsory detention occurs, and to explore potential pathways to prevent compulsory detention. A study exploring experiences of staff working in mental health services with people who have been involuntary admitted to hospital under the Mental Health Act was also conducted and is reported in a separate paper.

## Methods

### Design

A co-produced qualitative approach was used in this study, with a group with a range of relevant experience steering design, conduct and writing up throughout. Ethical approval to conduct the study was received from UCL Research Ethics Committee (Ref: 15249/002).

### Research team

The design, conduct and analysis of the study was co-produced by Lived Experience Researchers (LERs) who are members of the FINCH Co-Production Group, and by other members of the FINCH study research team. All LERs had personal experience of using mental health services and of compulsory detention, and/or of supporting and caring for a relative or friend who had experienced compulsory detention. The FINCH Co-Production Group, consisting of LERs, carers, clinicians and researchers (some with multiple roles), was established at the start of the FINCH study, and met fortnightly through the first year of the FINCH study to plan the current study and to develop the FINCH intervention and methods for our feasibility trial. The LERs received training in conducting online interviews, analysis and obtaining verbal informed consent. A monthly lived experience reflective space provided LERs with emotional support and space to discuss the research process and emotional impact during the study.

### Participants

Adults in England who had been detained under section 2 (mental health assessment) or section 3 (treatment for mental illness), of the Mental Health Act once or more in the last 5 years, were aged 18 years or over and had capacity to consent were eligible to take part in the study. Purposive sampling was used to ensure diversity regarding participants’ ethnicity, gender, age, self-reported diagnosis, geographical location, and number of times detained. We reviewed our sample during recruitment and implemented targeted strategies to ensure diversity. These included approaching community organisations working with Black and Minority Ethnic communities.

### Recruitment

We contacted several community organisations and national charities that support people with mental health problems, asking them to share the study poster with people accessing their organisation. We also asked the National Survivor User Network (NSUN), a network of individuals and user-led groups with lived experience of mental ill-health, distress, or trauma based in the UK, to disseminate the study adverts. We used Twitter to disseminate an invitation to the study through personal, study and institutional accounts, and asked organisations with Twitter accounts, Facebook and Instagram accounts to disseminate the study adverts. Potential participants contacted the research team by email. Researchers then checked eligibility, provided a participant information sheet, answered questions about the study, prior to booking interviews for those eligible and interested in taking part.

### Data collection

The interview topic guide was developed collaboratively by the Co-Production Group and the FINCH research team. We aimed to explore each participant’s most recent experience of being detained in hospital under section 2 or 3 of the Mental Health Act and of the support received following discharge, and to understand their views about what might have prevented them from being detained. The topic guide also asked participants’ views on the intervention being developed within the FINCH study, this further informed intervention development but is not reported in this paper. Interviews were conducted between September and December 2021. Please see Appendix A for the topic guide.

The interviews were conducted by five LERs with another FINCH researcher present to support recording and ensure the recording was securely stored in password protected university files. The interviews were conducted on Zoom or Teams videoconferencing platforms, and participants also had the option to phone in to Teams via a freephone number.

Verbal consent to take part in the interviews was recorded at the start of the interview. Socio-demographic information was collected via a secure online survey on the Opinio survey software. All interviews were recorded and were transcribed verbatim. All transcripts were then checked by the researchers and any identifying information was anonymised.

### Data analysis

We used Template Analysis [25], a form of thematic analysis [26] which involves the initial development of a coding template, based on a subset of transcripts, which is then applied to further transcripts and revised and refined as more data is analysed. This approach supports analysis by a group of researchers and ensures a focus on collaboratively defining meanings and structure of themes during the process of analysis [25]. Preliminary analysis of six transcripts was undertaken by six LERs to develop themes and subthemes as a basis for the first version of the analysis framework. Further transcripts were analysed by LERs and FINCH researchers, and an analysis meeting took place to revise the themes and subthemes in the analysis framework. The analysis framework was revised where new themes were added, or similar themes or subthemes merged. The remaining transcripts were then analysed using the revised framework. A final analysis meeting was held to further review the analysis framework to merge similar themes or subthemes, or add new themes identified.

### Reflexivity

The research team comprised of Lived Experience Researchers, with experience of using mental health services and of compulsory detention, and/or of supporting and caring for a relative or friend who has experienced compulsory detention, and other researchers included clinical academics and non-clinical researchers from a range of backgrounds, including a psychiatrist, psychologist, social worker, and an occupational therapist. The research team members are from diverse backgrounds in terms of ethnicity, age, and gender.

## Results

We recruited 20 people, of whom 11 (55%) were male and the most common age range was 25-29 years (35%). The most frequent group was Black/Black British (40%), with 30% White British, and Mixed/multiple ethnic groups (15%) the next most represented ethnic groups. Half of the sample (50%) lived in London, with 60% living alone. Seventy percent of the sample had been compulsory detained under the MHA more than once, and psychosis was the most common mental health diagnosis (35%). Table 1 presents more details of the participants’ demographic characteristics.

**Table 1.**
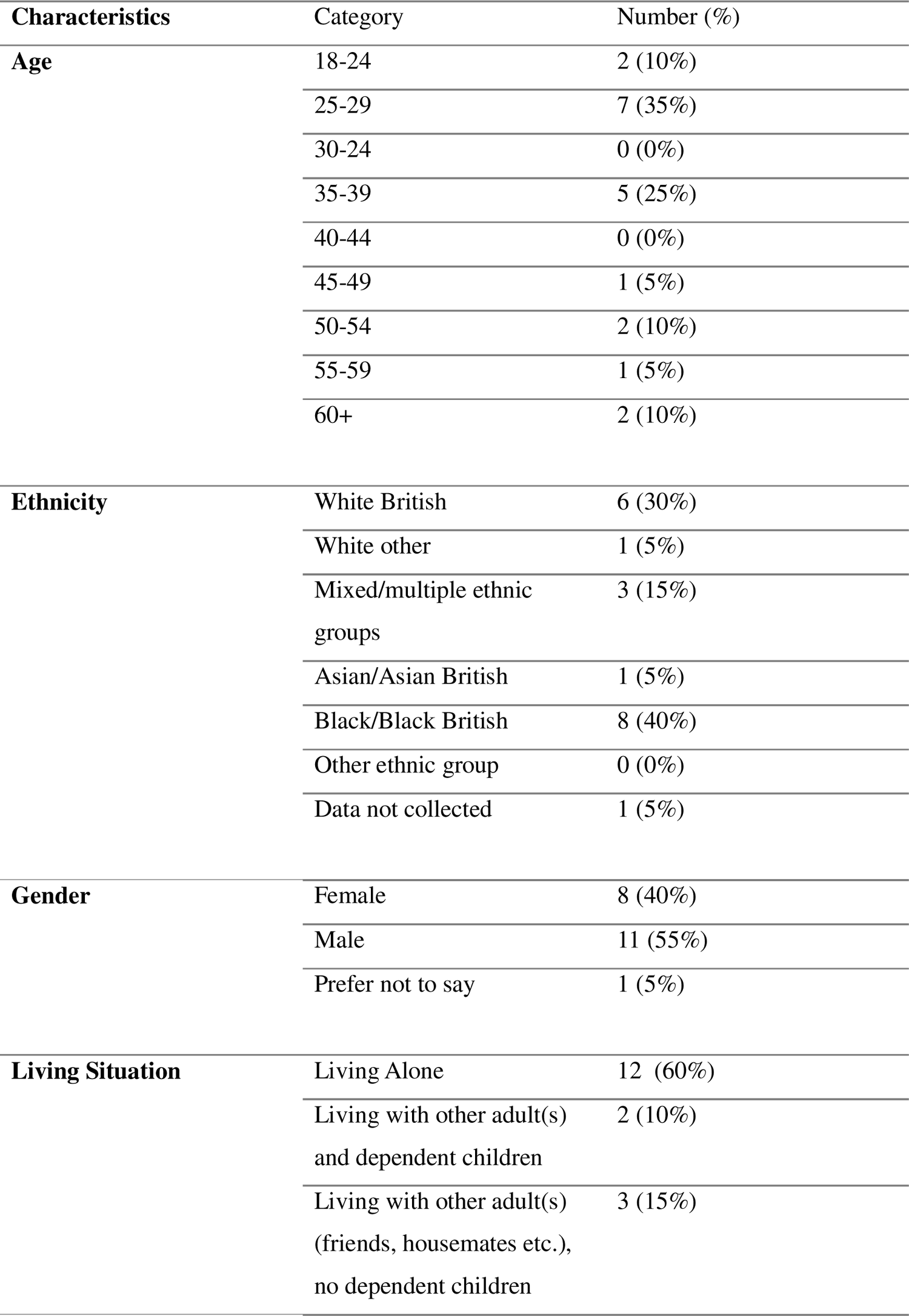
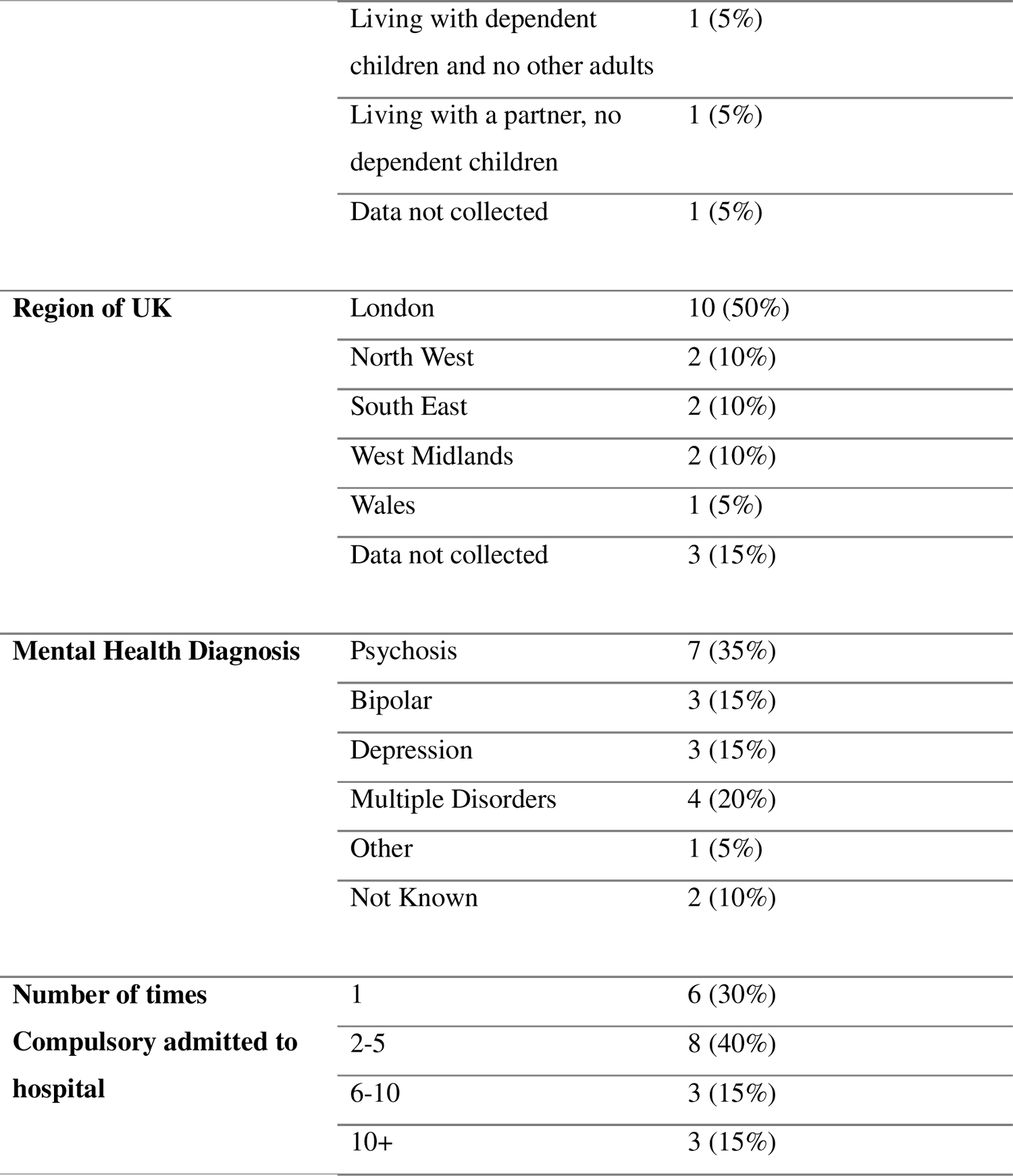
Characteristics of Participants (N = 20)

Three overarching themes were identified, and within them sub-themes that relate to factors found to increase the likelihood of being repeatedly detained in hospital under the Mental Health Act (MHA) and, suggestions for preventing this. Overarching themes related to the individual, family and support network, and improving service responses. A summary of each theme is outlined in Table 2.

**Table 2.** Overview of Themes and Sub-Themes.

| Theme | Sub-Themes |
| --- | --- |
| **Individual Factors increasing or reducing likelihood of being detained** | Life Stressors and Other Events<br>Attitudes to Own Mental Health & Self-Management Skills<br>Not Taking Prescribed Medication<br>Risk to Individual and Others |
| **Family and Support Network** | Attitude and Support from Family and Friends<br>Wider Social Networks as a Protective Factor to Maintain Mental Health |
| **Need for improvement in Service Responses** | Collaborative Care and Choice<br>Quality of Professional Support<br>Discriminatory Attitudes |

### Individual factors increasing or reducing likelihood of being detained

This theme encompasses factors relating to the participants’ own behaviour and feelings that they saw as contributing to becoming unwell and being detained in hospital under the MHA. Participants also suggested individual strategies that might help prevent future detentions under the MHA, and ways of supporting these.

#### Life Stressors and Other Events

Participants reported that recent and past events and current social stressors contributed to them being detained in hospital under the MHA. Some participants could see patterns to their detentions under the MHA over time, for example, seasonal patterns or the influence of recurring stressors were identified. Work-related stress, health concerns, illicit drug use, recent or past recurring traumatic experiences, bereavement, breakdown of relationships and an accumulation of problems with housing and family were reported as contributing factors:

> *“I was depressed about a lot of things, I had a lot of challenges facing me then, I lost a very very close friend of mine, he’s more like a brother to me, I lost him in a gun violence incident, it was really a traumatic experience for me….I was just really losing it”* [P14]

#### Attitudes to own Mental Health & Self-Management Skills

Some participants spoke of having a lack of awareness regarding the deterioration of their mental health and viewed an inability to mobilise self-management skills as contributing to eventual detention. For example, one participant stated that at the point of their mental health worsening:

> *“I had no consciousness to decipher my actions. I didn’t decipher my actions at that particular point. I was just freestyling.”* [P12].

Some also found it difficult subsequently to make sense of what happened at the time of them being detained, and therefore felt unable to know what to do differently to prevent them being detained in hospital under the MHA again:

> *“It is so traumatic being sectioned for anybody, and no one ever really talks about it after. It’s a really strange thing because you’re discharged and you’re back with your old team and it all just, sort of, disappears and you think, “Did I really just go through all that? What’s going on?”* [P4]

However, others described an increase in self-awareness regarding mental health subsequent to treatment in hospital:

> *“I was never a kind of self-awareness person. I always saw things from a distance and I never really gained a lot of self-awareness. But after that [creating a self-management plan] I had to become a more self-aware person.”* [P12].

Choosing to better manage their mental health post-discharge in order to prevent readmission was also reiterated by other participants:

> *“Yes. I think… I just need to work on myself and bring myself back home. That was just it for me. And since my last sectioning, I think I’ve been doing pretty well. I’ve been able to manage all those mental stresses that I’ve been having and managing my family”* [P19]

Some participants wanted to take action to support themselves on discharge to prevent further detentions under the MHA:

> *“actually… I really don’t wish.. to get in a situation that could get me sectioned again, it was really an awful time and that’s why I am very very intentional right now about managing my mental health.”* [P14]

#### Not Taking Prescribed Medication

Not taking prescribed medications was identified as a frequent factor leading to repeat detentions. Participants stopped taking medication for numerous reasons, including voices telling them to stop taking the medication; not liking the medication prescribed; being unable to resist a strong urge to stop medication; being unsure what the medication was; and feeling they did not need to take it.

> *“Well, there wasn’t any reasons for me to stop, I just felt I didn’t need them […] I didn’t really have a clear understanding of what the medication was. I just believed the medication was for my mental health, maybe to help me get calmer […] Yes. Maybe I realised, but then I didn’t just care.”* [P19]

Having a regular weekly routine, particularly regarding medication, was identified as a strategy to prevent future detentions.

> *“I think routine and a schedule to the week is always helpful so when it (medication) says evening, I take it in the early evening”* [P3]

#### Risk to Individual and Others

Participants reported engaging in behaviours, such as suicide attempts, which caused risk to themselves, then triggering a MHA assessment and detention. Others reported behaving violently as their mental health declined, leading to detention.

> *“So I just found myself really being this kind of violent, rowdy individual, fuming, and I had a lot of fury. It didn’t go well.”* [P10]

Illicit drug use and lack of self-care were other factor identified as leading to compulsory detention:

> *“I was not, you know, in a very good shape at all. I wasn’t taking care of myself. At that point in time, I hardly was eating. I was actually very violent and I resorted to heavy drug consumption.”* [P19]

### Family and Support Network

This theme describes how an individual’s family and support network could impact positively or negatively on service users’ mental state and functioning, potentially either contributing to repeat detentions or helping to stay out of hospital.

#### Attitude and Support from Family and Friends

Some participants described having disconnected and poor relationships with family and friends as contributing to repeat detentions. This included some describing feelings of rejection, lacking social support, and feeling misunderstood and misrepresented by their family.

> *“I think the issue is, once you’ve been labelled, you’re disbelieved. So if you’ve got family around you who are sabotaging you and not being very helpful, and don’t have a label, they’re automatically believed.”* [P7]

This participant further stated they had a particularly negative relationship with their mother:

> *“My mum has died now, but she was really, really abusive. But she was the one instigating it all. She was a little, old lady, and they believed what she had to say. So that’s quite difficult, I think. She’d be classed as the nearest relative.”* [P7]

In contrast, some participants described their family or friends as important in supporting them through their experience of mental ill-health. This included family or friends noticing changes in the participants’ behaviour and mental state, and encouraging them to seek help, intervening at the point of crisis, and supporting them through their mental health assessment by providing the individuals medical history to service staff.

> *“I was just all locked up in my thoughts, very distressed and I decided to end it and I was in the middle of trying to go ahead with the suicide when I had a friend come round and saw me in the process.”* [P15]

#### Wider Social Networks as a Protective Factor to Maintain Mental Health

Participants discussed how their wider social network had been beneficial in maintaining their mental health and thus preventing readmission. These included being in a positive employment role and engaging in external activities and community groups.

Engaging in local community groups was one example of how participants felt they could keep well and avoid readmission:

> *“I was connected to friends because I was told, “Do what you think can make you happy.” What could make me happy is playing football […] That really helped me because when I interacted with new friends and at least divert my energy to playing, it made me less distressed.”* [P11]

Workplace support and flexible working, which allowed the individual to maintain their employment, was also found helpful by some:

> *“I was encouraged by the fact that my employer decided not to fire me but, instead, decided to give me quite a relaxed timeline and timetable. My colleagues, also, were always there for me.”* [P14]

### Need for improvement in Service Responses

This theme encompassed problems in both inpatient and community services that may make compulsory detentions more likely to occur, and suggestions for improvements to address these. These challenges pertained to both inpatient detention and community-level care.

#### Collaborative Care and Choice

Several participants emphasised the importance of being given choice in treatment, and felt this was lacking in all settings, making it less likely that treatment would be accepted voluntarily either in hospital or in the community. Regarding inpatient admissions, many said that medication was the primary treatment delivered rather than alternatives being offered, and that choices regarding which drugs would be used were not offered:

> *“In hospital, they just gave me so many drugs, and I had no choice but to take them. They were forcibly given. I just complied, then, after that […] I only complied because if I didn’t, they’d have just kept injecting me with stuff. So I just ended up taking it.”* [P1]

There was also a lack of collaborative decision-making between service user and staff, for example in discussions at the point of detention:

> *“there’s this me versus them. There was no sense of collaboration, no sense of, ‘We’re here to help you.’ It was, ‘We’re here to detain you,’ […] That set the tone for the whole inpatient stay.”* [P1]

Some participants felt that they would have been less likely to be detained at their last admission if alternatives had been discussed:

> *“It would have been helpful because I’m not someone who likes hospitals very well. The smell around hospital makes me very uncomfortable. I would have appreciated other options apart from being sectioned.”* [P16]

#### Quality of Professional Support

Negative experiences of care from mental health services were described by several participants. Poor communication was a primary example, including staff not listening to the service user or their wider network.

> *“my community team didn’t listen to the third sector organisation that was supporting me.”* [P9]

Some participants believed that if clinicians valued their voice, compulsory detention would have probably have been avoided.

> *“So their whole assessment and decision making process really needs to be looked at. They ought to listen really to me and to the people who know me who I work with etc. because each time if they had done that there wouldn’t have been a hospitalisation, for the last three times.”* [P8]

Several participants also described staff as lacking interest in the individual service user; this included staff dismissing current care plans the service users wished to share with the team.

> *“I had all my recovery plans and my crisis plans in my bag, and they did not even want to read it…But nobody has been interested in the last, I do not know, five years or so.”* [P9]

Crisis plans and having a structured care plan was seen as something that could have prevented previous compulsory detentions.

> *“I think having a very structured comprehensive care plan in writing, accepting and going through it, and have, kind of, a crisis plan as well. Yes, I think, you know, I could have avoided many, many sections and many hospital interventions, because I could see the triggers, and I could see myself getting… I need to talk, and I need to, kind of, get a clear head, and avoid certain places or certain people.”* [P9]

Another example of poor quality support was the lack of information provided by staff to service users regarding medication.

> *“Well, there wasn’t any reasons for me to stop [taking medication], I just felt I didn’t need them. I didn’t really have a clear understanding of what the medication was. I just believed the medication was for my mental health, maybe to help me get calmer”* [P19]

Participants also discussed a lack of continuity of relationships with staff.

> *“It’s always different people, and then you just have to-even though they know the basics because they have your information, but you still end up having to repeat yourself again and again.”* [P20]

Many participants felt that being offered more post-discharge care, more accessible and available community support, alternatives to admission such as crisis houses, where they could feel safer and have more freedom, would have decreased the likelihood of them being compulsory detained on their last detention.

As well as in community settings, many participants felt that inpatient staff lacked interest in the service user at the point of discharge, including not providing a discharge or crisis plan, no information on medication withdrawal, and no acknowledgement of the individual’s external responsibilities.

> *“They wouldn’t let me look at the bus number. I had barely any money. They didn’t help with that. They were really unhelpful. So I left the hospital with about £10 in my pocket, I had no phone, and I was withdrawing from all sorts: benzodiazepines, antipsychotics, mood stabilisers. So I was in quite a bad shape. But no, they didn’t give me any plan or help, or anything.”* [P7]

#### Discriminatory Attitudes

Discriminatory attitudes among staff were described by one participant. This included discriminatory attitudes towards specific race or ethnicity and clinicians having pre-existing assumptions and/or stereotypes based on certain races/ethnic groups.

> *“I feel that it has a very strong element of racial discrimination there because the perception is, as a black woman at that, that I’m aggressive, that I’m difficult to deal with. And all of these things also have connotations with, “Black women act like that because they’ve got mental health difficulties.”* [P13]

Crisis plans and having a structured care plan was seen as something that could have prevented previous compulsory detentions.

> *“I think having a very structured comprehensive care plan in writing, accepting and going through it, and have, kind of, a crisis plan as well. Yes, I think, you know, I could have avoided many, many sections and many hospital interventions, because I could see the triggers, and I could see myself getting… I need to talk, and I need to, kind of, get a clear head, and avoid certain places or certain people.”* [P9]

## Discussion

### Main Findings

We identified three over-arching themes, encompassing potential contributing factors to being detained at service user, family and informal support network and mental health service levels, with participants putting forward practical suggestions for addressing these. The first theme “Individual factors increasing or reducing likelihood of being detained” reports external factors such as current and past life stressors and trauma, as well as internal factors such as individuals’ attitudes to and management of their mental health and decisions not to not take medication. Establishing routines including a medication regimen and self-management plans were reported as potentially preventing detention. The second theme “Family and Support Network” identified negative relationships with family as contributing factors to detention, and conversely having and keeping in contact with a supportive family, friend and wider network was seen as keeping people well and thus preventing detention. The third theme “Improving Service Responses” pinpoints limitations of services such as a lack of collaborative care and choice, poor quality of professional support, and discriminatory attitudes by staff which contribute to detention and the necessity for these to be improved to lessen the prospect of compulsory detention. Thus service user accounts suggest a variety of types of contributing factors to compulsory detention, with a range of potential strategies for addressing them, most of which seemed to the research team’s coproduction group potentially feasible approaches. Whilst three over-arching themes reflecting different domains were identified by the coding team, there were multiple connections between them, for example not taking medication in the context of lack of continuity of support and collaborative decision-making, or of lack of family support.

### Findings in the context of other studies

The focus of the current study on exploring service users views about what leads to compulsory admission is relatively novel: we are aware of little previous qualitative literature focusing on this particular question. An interview study in the UK exploring patients’ experiences of the assessment process identified lack of choice and of voice and involvement in decisions as central in these experiences [27]. Wormdahl et al. [22] carried out qualitative interviews and focus groups with patients and clinicians in Norway exploring pathways to involuntary admission: they identified a complex network of contributing factors, reflecting, as in our findings, both individual and service level difficulties, including living in deprived circumstances, discontinuing medication, and lack of responsiveness, collaboration and choice in mental health services. In a metasynthesis of the worldwide literature on involuntary admission, Akther et al, [4] found that patients’ lack of involvement in decision making and care planning at all stages in their care, and also reported that medication adherence was undermined by patients not having a good understanding of what medication was for or how it is supposed to work. Lack of choice and coercion in varying forms has been reported in other literature to be especially characteristic of involuntary admission, but to be a feature of relationships with services for many people in many settings [28, 29].

Our findings also cohere with some quantitative findings: for example, service user reports that lack of choice at various stage in the pathway to admission and of collaborative and supportive services fit with findings in the synthesis of quantitative literature [17] regarding associations between compulsory admission and poor quality community care and lack of admission alternatives. Participants in our study also suggested that collaborative planning of care, for example for crises, has potential to reduce admissions: this is in keeping with the findings from synthesising literature on prevention of compulsory admissions that interventions based on crisis planning are thus far the only approach supported by a substantial trial literature [14, 15]. A quantitative review of individual factors associated with compulsory admission [16] fits with some of the individual level factors identified by participants in our study including living in deprived circumstances, non-adherence to medication and behaviour becoming aggressive.

Participants from a wide range of ethnic groups across England took part in the study, and only one participant talked about experiencing discriminatory attitudes from staff which they perceived as contributing to them being detained under the MHA. This contrasted with findings from another qualitative study where people from a Black ethnic background in England were directly asked about racism and discrimination in the detention process, eliciting multiple reports of this [13]. Participants in our study did not report police involvement as a factor that contributed to their detention, whereas a review of quantitative studies of association found a strong association between police involvement in detention and involuntary care [16].

### Strengths and limitations

The current study had several strengths, including the novelty of the research question. There was considerable involvement from people with direct relevant personal experience who were involved as LERs in the planning, conducting and analysis of this study. Forty percent of the participants in this study were also from Black/Black British ethnic groups, whereas previous qualitative research [4] has been limited by the lack of participants from the most highly represented groups of those detained under the MHA. However, there were few accounts of experiences with the police, a potentially important omission, and our process for recruitment from a variety of non-health service sources may have missed some important perspectives. However limitations include having a heterogenous sample which limits the understanding of mechanisms and pathways for specific ethnic groups or other sub-groups of participants. Many participants also had difficulties recalling what happened during the MHA assessment process, and reflected relatively little on the discussions that took place at this time or what options were discussed during the assessment, during the interviews. Interviewing participants immediately after discharge may help support participants to recall the MHA assessment experience whereas participants in our sample had been detained up to 5 years ago. Furthermore, explicit questions about the cultural appropriateness of care or experiences of discrimination were not included in the interview topic guide as our approach was to ask very broad open questions: thus we may well not have elicited all that respondents could have said about this. The use of multiple interviewers and a large analysis team with a wide range of experiences and characteristics may have enhanced the validity of themes, identified from multiple perspectives, but may also have increased variability in how interviews and analytic coding were conducted.

### Clinical and Policy Implications

Participants’ views of what might contribute to and prevent compulsory detention were often practical in nature and fitted well with the published evidence, where available, supporting several potential pathways to preventing detentions and repeat detentions. At the individual level, service users’ views supported the potential value of supported self-management and of interventions to alleviate problematic social circumstances [30, 31], and the importance of clinicians taking into account advanced choices, also supported by national policy in England [32].

Our results indicated that family involvement may have value in preventing detention, but could also be experienced as unhelpful, suggesting that involvement should be patient-led where possible and include educating families and the offer of evidence-based family interventions. Interventions that aim to involve family and friends in care such as Open Dialogue, may support patient-led involvement, although evidence is not definitive currently [33]. Family involvement is among the aspects of care that are the focus of proposals in the government White Paper in England and Wales on Reforming the Mental Health Act [32]. This also proposes more detailed risk assessment with a higher threshold for compulsory detention, whereby the individual would need to present with a substantial likelihood of significant harm, and a statutory obligation for detained service users to have a detailed and collaborative care and treatment plan [32]. These are all proposals that are in keeping with our findings, although as yet law as of March 2024.

At service level, the value of talking to service users about what is likely to be helpful is reflected in clear suggestions for improvements at a range of levels, including crisis planning, supported self-management, and collaborative decision making, all of which also have quantitative evidence of effectiveness in general, with crisis plans being the only intervention with substantial evidence of effectiveness in preventing detentions [14, 15, 31]. Our findings also suggest that interventions to improve alliance with staff, service user voice and collaborative decision making at all stages are potentially valuable in preventing detentions [34]. Participants highlighted a lack of voice or collaboration in the assessment process, and this links with findings in a previous study [30] and highlights the need for initiatives to improve the MHA process. It is also important for staff to explain their role and be considerate of endings/continuity of care and of collaborative care. Discharge following a compulsory admission emerges as a crucial point, when formulation of clear care plans, education on medication and self-management, patient-led engagement with families and friends and consideration of social support and mental health support in community context may prevent further detentions under MHA. At a service system level, provision of community alternatives when in crisis [35] and focusing resources on long term, high quality and individualised community care in which service users are supported to participate in decisions and provided a broad range of help with supported self-management, medication management and social difficulties was supported by our findings as a potential means of reducing compulsory admission. A single participant referred to discrimination, but many participants felt disempowered and lacking a voice in the care, an experience to which racial inequalities may also have been relevant [36].

### Further Research

Our findings supported a range of potential strategies to reduce detention, with more choice, better support and more effective care planning all approaches that were supported both by our findings and other qualitative and quantitative studies in related areas. Evidence evaluating the effectiveness of interventions to reduce detention, or assessing the impact of different community support arrangements on detentions remains very limited [14, 17], so that both trials of novel interventions to prevent detention among people at high risk and naturalistic evaluations of the relationship between community and crisis care arrangements and detentions are potentially of value. Although we ensured we had a diverse sample, including groups at high risk of compulsory detention, we did not specifically explore relationships between experiences of racism and discrimination or cultural identity and risk of detention; a more in-depth approach is likely to be beneficial.

## Conclusion

This study has identified complex interacting factors that may contribute to people being detained in hospital under the MHA, and that strategies for improved services, including more collaborative care in addition to person led family involvement may prevent further detentions. Increasing collaborative care, self-management and family and wider support can reduce further detentions. Interventions specifically aiming to reduce further detentions and focusing on these aspects of care need to be developed and tested.

## Data Availability

All data produced in the present work are contained in the manuscript

## Author Abbreviations

Ariana Kular (AK); Brynmor Lloyd-Evans (BLE); Cady Stone (CS) Fiona Lobban (FL); Henrietta Mbeah-Bankas (HMB); Jackie Hardy (JH); Janet Seale (JS); Jordan Parkinson (JP); Kathleen Lindsay Fraser (KLF); Lisa Wood (LW); Lizzie Mitchell (LM); Mark Keith Holden (MKH); Mary Birken (MB); Patrick Nyikavaranda (PN); Sonia Johnson (SJ); Tyler Elliott (TE); Valerie Christina White (VCW); Zishi Li (ZI)

## Ethics Declarations

### Ethics approval and consent to participate

Full Health Research Authority (HRA) and NHS Research Ethics Committee (REC) approval has been granted by the London - Bromley Research Ethics Committee (IRAS: 300671; Protocol number: 143180; REC reference: 21/LO/0734). Participants will be required to provide informed consent before inclusion into the study.

### Consent for publication

Not applicable.

## References

1. Hatfield B, & Antcliff V. (2001) Detention under the Mental Health Act: Balancing rights, risks and needs for services. Journal of Social Welfare and Family Law, 23(2), 135–153. 10.1080/01418030122309

2. NHS Digital (2022a) Mental Health Act Statistics, Annual Figures, 2021-22. Retrieved 26 January 2023, from https://digital.nhs.uk/data-and-information/publications/statistical/mental-health-act-statistics-annual-figures/2021-22-annual-figures

3. Sheridan Rains L, Zenina T, Dias MC, Jones R, Jeffreys S, Branthonne-Foster S, et al. (2019) Variations in patterns of involuntary hospitalisation and in legal frameworks: an international comparative study. The Lancet Psychiatry, 6(5), 403–17. 10.1016/S2215-0366(19)30090-2

4. Akther SF, Molyneaux E, Stuart R, Johnson S, Simpson A, Oram S. (2019) Patients’ Experiences of Assessment and Detention under Mental Health legislation: Systematic Review and Qualitative meta-synthesis. BJPsych Open, 5(3). 10.1192/bjo.2019.19

5. Stuart R, Akther SF, Machin K, Persaud K, Simpson A, Johnson S, et al. (2020) Carers’ experiences of involuntary admission under mental health legislation: systematic review and qualitative meta-synthesis. BJPsych Open, 6(2). 10.1192/bjo.2019.101

6. Department of Health and Social Care (2018) Modernising the Mental Health Act – final report from the Independent Review. London: DHSC.

7. Aluh DO, Ayilara O, Onu JU et al. (2022) Experiences and perceptions of coercive practices in mental health care among service users in Nigeria: a qualitative study. Int J Ment Health Syst, 16, 54. 10.1186/s13033-022-00565-4

8. Shozi Z, Saloojee, S, & Mashaphu S. (2023) Experiences of coercion amongst involuntary mental health care users in KwaZulu-Natal, South Africa. Frontiers in psychiatry, 14, 1113821. 10.3389/fpsyt.2023.1113821

9. National Institute of Clinical Excellence (2019). Service user experience in adult mental health services. Retrieved 28 July 2023, from https://www.nice.org.uk/guidance/qs14

10. NHS (2022) Mental Health Act. https://www.nhs.uk/mental-health/social-care-and-your-rights/mental-health-and-the-law/mental-health-act/ Accessed 12 Oct 2023

11. Barnett P, Mackay E, Matthews H, Gate R, Greenwood H, Ariyo K, Bhui K, Halvorsrud K, Pilling S, & Smith S. (2019) Ethnic variations in compulsory detention under the Mental Health Act: a systematic review and meta-analysis of international data. Lancet Psychiatry, 6(4), 305–317. 10.1016/S2215-0366(19)30027-6

12. NHS Digital (2022b). Detentions under the Mental Health Act. Retrieved 19 May 2023, from https://www.ethnicity-facts-figures.service.gov.uk/health/mental-health/detentions-under-the-mental-health-act/latest#by-ethnicity-5-ethnic-groups

13. Solanki J, Wood L, & McPherson S. (2023) Experiences of adults from a Black ethnic background detained as inpatients under the Mental Health Act (1983). Psychiatric Rehabilitation Journal, 46(1), 14–20. 10.1037/prj0000537

14. Bone JK, McCloud T, Scott HR, Machin K, Markham S, Persaud K, Johnson S, & Lloyd-Evans B. (2019) Psychosocial Interventions to Reduce Compulsory Psychiatric Admissions: A Rapid Evidence Synthesis. EClinicalMedicine, 10, 58–67. 10.1016/j.eclinm.2019.03.017

15. Molyneaux E, Turner A, Candy B, Landau S, Johnson S, & Lloyd-Evans, B. (2019). Crisis-planning interventions for people with psychotic illness or bipolar disorder: systematic review and metaanalyses. BJPsych Open, 5:4, 10.1192/bjo.2019.28

16. Walker S, MacKay E, Barnett P et al. (2019) Clinical and social factors associated with increased risk for involuntary psychiatric hospitalisation: a systematic review, meta-analysis, and narrative synthesis. Lancet Psychiatry, 6: 1039–53. 10.1016/S2215-0366(19)30406-7

17. Aluh DO, Aigbogun O, Ukoha-Kalu BO, Silva M, Grigaitė U, Pedrosa B, Santos-Dias M, Cardoso G, Caldas-de-Almeida JM. (2023) Beyond Patient Characteristics: A Narrative Review of Contextual Factors Influencing Involuntary Admissions in Mental Health Care. Healthcare, 11(14):1986. 10.3390/healthcare11141986

18. de Jong MH, Wierdsma AI, Zoeteman J, van Boeijen CA, Van Gool AR, Mulder CL. (2021) Risk factors for repeated emergency compulsory psychiatric admissions. BJPsych Open, 7(1):e19. 10.1192/bjo.2020.153

19. Sheridan Rains L, Weich S, Maddock C, Smith S, Keown P, Crepaz-Keay D, Singh SP, Jones R, Kirkbride J, Millett L, Lyons N & Lloyd-Evans, B. (2020) Understanding increasing rates of psychiatric hospital detentions in England: development and preliminary testing of an explanatory model. BJPsych Open, 6(5), e88, 10.1192/bjo.2020.64

20. Freitas DF, Walker S, Nyikavaranda P, et al. (2023) Ethnic inequalities in involuntary admission under the Mental Health Act: an exploration of mediation effects of clinical care prior to the first admission. The British Journal of Psychiatry, 222(1):27–36. 10.1192/bjp.2022.141

21. Sugiura K, Pertega E, Holmberg C. (2020) Experiences of involuntary psychiatric admission decision-making: a systematic review and meta-synthesis of the perspectives of service users, informal carers, and professionals. Int J Law Psychiatry, 73:101645. 10.1016/j.ijlp.2020.101645

22. Wormdahl I, Husum TL, Kjus SHH, Rugkåsa J, Hatling T, & Rise MB. (2021) Between No Help and Coercion: Toward Referral to Involuntary Psychiatric Admission. A Qualitative Interview Study of Stakeholders’ Perspectives. Frontiers in Psychiatry, 12.

23. Lay B, Kawohl W, Rössler W. (2018) Outcomes of a psycho-education and monitoring programme to prevent compulsory admission to psychiatric inpatient care: a randomised controlled trial. Psychological Medicine, 48(5):849–860. 10.1017/S0033291717002239

24. Johnson, S., Birken, M., Nyikavaranda, P., Kular, A., Gafoor, R., Parkinson, J., et al (2024) A crisis planning and monitoring intervention to reduce compulsory hospital readmissions (FINCH study): protocol for a randomised controlled feasibility study. Pilot and Feasibility Studies, 10, 35. 10.1186/s40814-024-01453-z

25. King N & Brooks JM. (2017) Template Analysis for Business and Management Students. London: Sage

26. Braun V & Clark V. (2019) Reflecting on reflexive thematic analysis. Qualitative Research in Sport, Exercise and Health, 11:4, 589–597, 10.1080/2159676X.2019.1628806

27. Blakley L, Asher C, Etherington A, Maher J, Wadey E, Walsh V, Walker S. (2022) ’Waiting for the verdict’: the experience of being assessed under the Mental Health Act. J Ment Health, 31(2), 212–219. 10.1080/09638237.2021.1922624.

28. Hoge SK, Lidz CW, Eisenberg M, et al. (1997) Perceptions of coercion in the admission of voluntary and involuntary psychiatric patients. Int J Law Psychiatry, 20(2):167–181. 10.1016/s0160-2527(97)00001-0

29. Szmukler G & Appelbaum PS. (2008) Treatment pressures, leverage, coercion, and compulsion in mental health care, Journal of Mental Health, 17:3, 233–244, 10.1080/09638230802052203

30. Barnett P, Steare T, Dedat Z et al. (2022) Interventions to improve social circumstances of people with mental health conditions: a rapid evidence synthesis. BMC Psychiatry, 22, 302. 10.1186/s12888-022-03864-9

31. Lean M, Fornells-Ambrojo M, Milton A, Lloyd-Evans B, Harrison-Stewart B, Yesufu-Udechuku, A, & Johnson S. (2019) Self-management interventions for people with severe mental illness: systematic review and meta-analysis. The British Journal of Psychiatry, 214:5, 260–268, 10.1192/bjp.2019.54

32. Department of Health and Social Care and Ministry of Justice (2022) Draft Mental Health Bill. Retrieved 15 Nov 2023, from https://www.gov.uk/government/publications/draft-mental-health-bill-2022

33. Pilling S, Clarke K, Parker G, et al. (2022) Open Dialogue compared to treatment as usual for adults experiencing a mental health crisis: Protocol for the ODDESSI multi-site cluster randomised controlled trial. Contemp Clin Trials, 113:106664. 10.1016/j.cct.2021.106664

34. Bowers L, James K, Quirk A, et al.(2015) Reducing conflict and containment rates on acute psychiatric wards: The Safewards cluster 24andomized controlled trial. Int J Nurs Stud., 52(9):1412–1422. 10.1016/j.ijnurstu.2015.05.001

35. Dalton-Locke C, Johnson S, Harju-Seppänen J, et al. (2021) Emerging models and trends in mental health crisis care in England: a national investigation of crisis care systems. BMC health services research, 21(1), 1174. 10.1186/s12913-021-07181-x

36. Smith SM, Kheri A, Ariyo K, Gilbert S, Salla A, Lingiah T, Taylor C, & Edge D. (2023) The Patient and Carer Race Equality Framework: a model to reduce mental health inequity in England and Wales. Frontiers in psychiatry, 14, 10.3389/fpsyt.2023.1053502

