## Supplemental Table 1 for "Exploring pathways to compulsory detention in psychiatric hospital and ways to prevent repeat detentions; Service user perspectives"

| Supplemental Table1: Additional Quotes | | |
|---|---|---|
| **Theme** | **Individual Factors Contributing to Detention or Potentially Preventing Detention** | |
| **Sub Themes:** | **Life Stressors and Other Events** | **Attitudes to Own Mental Health & Self-Management Skills** |
| **Additional Quotes** | "*Yeah, a lot of things were going on around me then. A lot of situations were not too good, a lot of things happening negatively and that was affecting me."* [P15]<br><br>"*I think I had undergone great stresses. I had lost my job and I was in great disarray. It led to me being really sectioned*." [P18]<br><br>"*Every single time I've been sectioned, there are always stresses and triggers, always thinking about past things that I start to remember. And then I get into it more and more, stop taking my medication, start saying doing things which is not normal. There are a lot of things*." [P20] | "*I wasn't taking care of myself, you know, personal hygiene. I just left it all out*." [P17]<br><br>"*I was feeling good hope at some point before I thought, "I'm good," and decided not to see my therapist or take my meds. I just decided to be an animal, and I really ended up being dirty, so to speak*." [P10] |

| **THEME** | **Individual** | |
|---|---|---|
| **Sub Themes:** | **Not Taking Prescribed Medication** | **Risk to Individual and Others** |
| **Additional Quotes** | "*I find the urge to stop taking medication becomes too great, if I stop taking medication I need to be in hospital."* [P3]<br><br>*"I was happy to take some meds but I wasn't happy to take the medication they wanted to give me which was an antipsychotic. And so they sectioned me, and I remained under section until the following November, so that was a year*." [P1]<br><br>"*Yes. But I am not very keen on medication, so I take the lowest possible medication that will manage my condition or my stress… I take it when I feel I need it to*." [P9] | "*My mental health really got really bad and was deteriorating. I was consuming a lot of drugs, drugs and alcohol. I think it actually started to deteriorate, my mental health, and I was sectioned."* [P17]<br><br>"*It's things I've said and done in the past which is why I was sectioned, talking about things which my family members are saying not making any sense, but they're worried about me, that I could hurt myself, hurt others."* [P20]<br><br>"*Well, the first one (*hospital admission under MHA*) was in response to a serious suicide attempt… The others (*hospital admissions under MHA*) were basically disproportionate risk aversion on the part of service."* [P8] |

| Theme | Family and Support Network | |
|---|---|---|
| **Sub Themes:** | **Attitude and Support from Family and Friends** | **Wider Social Networks as a Protective Factor to Maintain Mental Health** |
| **Additional Quotes** | "*They were big friends of mine* [ work-friends] *I think they made a difference because they were there to provide my history to the medics, and even as I went through therapy, they gave me a realistic aspect of life."* [P10]<br><br>"*By chance I finally made a friend. She was the one who noticed these things about me. So, she was the one who encouraged me to seek medications. Although it wasn't really what I wanted to do."* [P16].<br><br>"*…I think my family were doing the best they can. My mum and my [brother and sister], they can't do any more than that, and they were worried about me […] They wish they're not having to make these phone calls and have people come in and section me, but it was either that or me doing something to myself or family members. So no, I don't blame them."* [P20] | "*What could make me happy is playing football. So I was connected to some friends …. That really helped me because when I interacted with new friends and at least divert my energy to playing, it made me less distressed*." [P11]<br><br>"*Some members of my church group were always coming around to the house and trying to engage me in activities, trying to get me to come out and back to my normal self, that's my youth group in the church*." [P14] |

| Theme | Improving Service Responses | | |
|---|---|---|---|
| **Sub Themes:** | **Collaborative Care and Choice** | **Quality of Professional Support** | **Discriminatory Attitudes** |
| **Additional Quotes** | "*I feel like the key word there is choice… if any service delivery is truly person-centred, then it needs to incorporate choice*." [P13] | "*But one of the things that I think contributed to my problems is the way the support worker just left a voicemail saying, 'Okay, I'm not working for you anymore now. That's it,' and not even asking me what I think about that..just making that decision. That didn't help, just basically being left by myself*." [P20]<br><br>"*the first time I was sectioned, it's like I had support for three years at a centre, so I was getting support. I had a support worker. I was going into sessions that the mental health clinic was doing. I was taking part in a lot of things, like men's groups and stuff like that…"* [P20] | "*I remember vividly, there was another person on the ward who came into my room and, like, we were doing colouring and she said to me that the staff had actually told her to stay away from me because I was trouble*." [P13]<br><br>"*There was sort of, it was like the head of the ward. It was a [man from an African country] and I've got [same African country] origins. So he could tell from my name where my heritage is. He was quite nice to me. He was actually very nice to me, yeah. So apart from the discrimination by other staff members, there were two other staff members that were actually quite pleasant to me*." [P13] |
