## Appendix A for "Exploring pathways to compulsory detention in psychiatric hospital and ways to prevent repeat detentions; Service user perspectives"

**Study: Exploring pathways to detention in psychiatric hospital and ways to prevent repeat detentions: a qualitative study**

**Interview Topic Guide for online semi-structured interviews with People with experience of being involuntarily admitted under the Mental Health Act**

This topic guide will be discussed with the Co-Production Group (consisting of researchers, clinicians and people with relevant lived experience) and piloted with at least three service users, after which some amendments to wording may be made.

**Introduction:**

**We are a research team, including mental health staff and people who have personal experience of using mental health services, and people who have caring experience. We are carrying out a study in which we are trying to develop a new type of support to help people who have been “sectioned” reduce their likelihood of being “sectioned” again in the future.**

**To help us develop this new type of support, we are trying to find out more about people’s experience of what led to them getting “sectioned”, and what you think could be helpful in stopping this from happening again.**

**We also want to ask you about our ideas for the new type of support and get your thoughts on how helpful and appealing it sounds, so we can try to make it as relevant and useful as possible.**

**We are aware that some of the questions are about difficult times in your life and could be upsetting – please only share what you feel comfortable talking about. We can take breaks during the interview so please let me know if you’d like to stop at any time.**

1. **When was the last time you were “sectioned in hospital”?**

**Can you please describe what happened in the run-up to being sectioned?**

*Prompt for: whether they think any stresses in their lives or any recent events that were out of the ordinary, had treatment and contact with services changed in any way, do they think their mental health was deteriorating? If they thought their mental health was deteriorating, did they know what to do? Also prompt for what happened on the day of the Mental Health Act assessment and on the days leading up to it, and what discussion they remember about options at this assessment or just before.*

2. **Do you think there is anything that could have made getting sectioned (again) less likely?**

*Prompt for: anything staff could have done, whether it would have helped to be in contact with different services, anything the person could have done themselves, anything family and friends could have done. Also, what kind of follow-up did they have after the last admission, and how was this? Did they drop out of care, and if so, what was done and could it have been different? Did they stop taking meds – if so, why did they stop taking meds, did they understand why it was recommended they take them? Were they having problems with the meds, and did they know how to get them reviewed? what was done after they stopped taking meds, and could it have been different? Ask about their care plan and risk management plan and whether they led the discussion on this and who was in charge of managing this in the community e.g. care co-ordinator etc.*

3. **Looking back to previous times you have been sectioned, did that follow a similar pattern? If not, what was different on past occasions?**

*Probe for any other risk factors, things that could be changed, different patterns of ending up back on section. Discuss some of the above without being leading.*

4. **When you last left hospital, did you have any kind of plan for staying well?**

**Were you given any guidance about keeping an eye on your mental health?**

**Did you know what to do if things got worse with your mental health?**

*Prompt for any kind of crisis plan, relapse prevention plan, risk management plan, community support,* *self-management plan, how they understand it and what they feel is being achieved. (Good to find out if a care, crisis plan, relapse plan was monitored or just produced without any follow up.)*

**Then: researcher will give a brief explanation of the intervention**:

The aim of the new type of support is to help prevent people who have been “sectioned” (admitted to hospital under the Mental Health Act”) from being sectioned again. First of all, people will be offered around 4 sessions with a clinical psychologist or another experienced therapist. These sessions will begin on the ward, or just after discharge. They will involve looking at what might have led to getting “sectioned” and what could be done to prevent this happening again, including both what the person could do themselves, and what their family and friends and staff working with them could do. Then there will be monthly phone or video calls for a year afterwards to check in and offer support in following through on plans to avoid being sectioned. Calls will include checking in about whether you have any concerns about your mental health or keeping out of hospital, and discussing what you could do if you are concerned.

5. **Does this sound helpful to you? Why / why not?**

*Prompt for positive and negative aspects, anything that could be changed or added. Include Video call or phone.* Do you feel this this approach could be helpful? for you and for others in a similar situation?

6. **Do you have any other ideas for this new type of support?**

*Prompt for understanding crisis and the factors that might lead to you getting unwell or ending up in hospital, understanding how services work and how to get the help you need, understanding more about mental health, looking at how family and friends can best provide support, looking at how staff can best provide support, monitoring warning signs. Getting people involved in decisions about their treatment, help them keep to medication plans. Understanding how your background (e.g. ethnic group, religion, sexuality) might affect the support you need. Community support, peer support, GP* *Etc.*

7. **With this new type of support, there are two options for how the personal mental health worker delivering the intervention might communicate with the person’s care team.**

**The first option is that the personal mental health worker providing the new type of support could be very separate from the person's care team and leave it up to the person how much they involve their care team in the crisis planning and monitoring.**

**The second option is that the mental health worker delivering the new support could communicate and link up more proactively with the person's care team, to share information and make arrangements for crisis support.**

**Which of these approaches do you think might be better and why?**

*Prompt for: Why do you think either option is better? / Would you prefer your care team to know about your involvement in a crisis-planning study, or not? Why? / What are the advantages and disadvantages of linking up the person delivering a crisis-planning intervention with someone’s care team?*

8. **Once the new type of support is ready, the next step is to run a study to encourage people to try it out.**

**What do you think would get people who are in hospital under a “section” interested in receiving this new kind of support?**

**What might encourage people to take part in a study to test the new kind of support?**

*Prompt for:* *What do you think might influence whether participants want to be in this study or not? / What might make it difficult for people to take part? / What might make it easier for people to take part? / How can we make it more interesting and appealing for potential participants?*
